# The indirect relationship between sleep and cognition in the PREVENT Cohort: Identifying targets for intervention

**DOI:** 10.1101/2023.05.18.23290160

**Authors:** Benjamin Tari, Michael Ben Yehuda, Axel Anders Stefan Laurell, Karen Ritchie, Yves Dauvilliers, Craig Ritchie, Brian Lawlor, Lorina Naci, Graciela Muniz Terrera, Paresh A. Malhotra, Tam Watermeyer, Robert Dudas, Benjamin R. Underwood, John T. O’Brien, Vanessa Raymont, Ivan Koychev

## Abstract

**Introduction:** As the global population ages, the economic, societal, and personal burdens associated with worsening cognition and dementia onset are growing. It is therefore becoming ever more critical to understand the factors associated with cognitive decline. One such factor is sleep. Adequate sleep has been shown to maintain cognitive function and protect against the onset of chronic disease, whereas sleep deprivation has been linked to cognitive impairment and the onset of depression and dementia.

**Objectives:** Here, we aim to identify and explore mechanistic links between several sleep parameters, depressive symptoms and cognition in a cohort of middle-aged adults.

**Methods:** We investigated data from the PREVENT dementia programme via structural equation modelling to illustrate links between predictor variables, moderator variables, and two cognitive constructs (i.e., Executive Function and Memory).

**Results:** Our model demonstrated that sleep quality, and total hours of sleep were related to participants’ depressive symptoms, and that, participant apathy was related to higher scores on the Epworth Sleepiness and Lausanne NoSAS Scales. Subsequently, depressive symptoms, but not sleep or apathy ratings, were associated with Executive Function.

**Conclusions:** We provide evidence for an indirect relationship between sleep and cognition mediated by depressive symptoms in a middle-aged population. Our results provide a base from which cognition, dementia onset, and potential points of intervention, may be better understood.

## 1.0 Introduction

As the global population grows older (United Nations, 2020) it has become increasingly important to understand the factors which contribute to healthy aging. The normal aging process impacts all physical and behavioural functions, including cognition (Harada et al., 2013). Broadly, cognition encompasses the crucial day-to-day abilities necessary to correctly respond to one’s environment and includes components such as stimulus processing, memory (Hadara et al., 2013), and higher-order executive functions (see Diamond, 2013). The trajectory of cognitive performance over time follows an inverted U. That is, cognition develops in childhood and adolescence, peaks during adulthood, and most domains begin to steadily decline after middle-age (Krivanek et al., 2021). Protecting cognition has garnered increased attention due to the massive economic and social burdens associated with its unhealthy decline and the onset of dementia (Alzheimer’s Association, 2021). A myriad of factors may influence the rate at which cognition declines and the likelihood of developing dementia, including sleep patterns (Scullin and Bliwise, 2015).

Adequate sleep is vital for the maintenance of cognitive performance (Dzierzewski et al., 2018; Matricciani et al., 2019) and appears to be critical for the clearance of beta-amyloid (Aβ) protein (see Wang and Holtzman, 2020). Regarding the latter, self-reported poor sleep quality has been shown to be related to higher Aβ concentrations in middle-aged and older adults (Spira et al., 2013; Sprecher et al., 2015; but see Gabelle et al., 2019^1^) and chronic sleep deprivation has resulted in greater accumulation of this neurotoxic protein (Tabuchi et al., 2015). More recently, Shokri-Kojori et al. (2018) demonstrated that acute sleep deprivation (i.e., 1 night) yielded increased Aβ concentrations in the hippocampus and thalamus of individuals aged between 22 and 72 years. These results may be related to decreased clearance of Aβ contingent upon adequate glymphatic function (e.g., Xie et al., 2013) and/or γ-oscillations during rapid eye-movement sleep (Aron and Yankner, 2016). Alternatively, results may reflect increased synthesis of Aβ in response to the lack of sleep (Castellano et al., 2011). These are important results as they demonstrate a potential link between acute and chronic sleep disruptions and the development of Alzheimer’s disease pathologies (Ju et al., 2013, 2014; Wang and Holtzman, 2020). Moreover, studies of sleep deprivation demonstrate a general worsening of cognition (for review see Killgore, 2010). For example, Lo and colleagues (2016) found that acute partial (i.e., 5 hours sleep for 7 nights) and total (i.e., 1 night) sleep deprivation contributes to the formation of false memories, and Gevers et al. (2015) demonstrate a general slowing of Stroop task reaction times (RT). Sleep deprivation is thought to impair the ability of the brain to consolidate memories (Yoo et al., 2007) and reduce the availability of the resources necessary for adequate stimulus processing and top-down executive control (e.g., Botvinick et al., 2001). Similarly, too much sleep (e.g., 10 or more hours) has been found to a risk factor for the development of global cognitive decline (Ma et al., 2020) and dementia onset (Cavaillès et al., 2022a). The mechanisms underlying the relationship between cognitive decline and sleep durations are not completely clear, but biological factors including elevated inflammation (Patel et al., 2009) and thinning in executive brain regions (Spira et al., 2016) have been proposed. Optimal health is likely supported by approximately 7 hours of sleep per night (Watson et al., 2015); however, overall sleep quality (i.e., related to factors such as sleep disturbances, trouble falling asleep, long waketime after sleep onset) is also related to global cognition. In a recent systematic review, Casagrande et al. (2022) identified that a greater frequency of sleep disturbance is associated with impaired cognition. Moreover, individuals who regularly sleep poorly are at a higher risk for developing symptoms of depression (Riemann et al., 2020) and dementia (Sabia et al., 2021). The mechanism(s) by which sleep influences the onset of depression are unclear. However, evidence indicates that poorer sleep quality may disrupt neural plasticity and synaptic health within the brain’s emotion processing regions (Disner et al., 2011; Riemann et al., 2020).

It is important to note that the relationship between sleep disruption and depressive symptoms is bi-directional. Symptoms of depression include poorer sleep quality, as well as increased fatigue, diminished concentration, decreased ability to make decisions, low mood, and apathy (Blazer, 2003). Importantly, and perhaps in conjunction with poorer sleep quality (Jaussent et al., 2011), depressive symptoms impair cognition (Varghese et al., 2022) and foster a greater likelihood for dementia development (Kessing, 2012; Cavaillès et al., 2022b). For example, Lindert et al. (2021) demonstrated that longitudinally, higher scores on the Centre for Epidemiologic Studies Depression (CES-D) scale were positively related to worse scores on measures of episodic memory and executive function. This relationship has been demonstrated across the lifespan (see also Dotson et al., 2020) and has been attributed to elevated cortisol concentrations (Sapolsky, 2000) and increased neurotoxicity due to increased inflammation (Furtado and Katzman, 2015). Taken together, the current research landscape provides evidence for the links between sleep, depression and cognition. Importantly, however, the mechanistic nature of these associations remains poorly understood, especially in a mid-age population. Here, the PREVENT cohort – composed of individuals between the ages of 40 and 59 – was used to examine the relationship between sleep, depression symptomology and cognition prior to any dementia diagnosis. We hypothesised that measures of sleep quality and quantity would be related to cognitive performance and it may be that this association is explained via an indirect association with depression or its symptoms.

## 2.0 Materials and Methods

### 2.1 Participants

700 individuals (age range 40 – 59) from the PREVENT dementia programme (Ritchie et al., 2013) were included in this investigation. All participants self-reported being cognitively healthy at the time of collection. We note that a minority of participants reported a current diagnosis of depression (n = 23), sleep disorder (n = 93), anxiety disorder (n = 60), mood disorder (n = 24), psychotic disorder (n = 2), alcoholism (n = 3) and drug misuse (n = 2). We note that the data used in these analyses is secondary data where ethical approval has been obtained by the source cohort (i.e., PREVENT).

### 2.2 Cognitive Assessments

Participants’ cognitive function was assessed via the COGNITO battery and tests included measures of executive function (i.e., Stroop colour, word, and interference tasks), and memory (COGNITO Tasks 8 – Articulation and Immediate Recall – and 17 – Delayed Recall of Names). These tasks were selected due to their sensitivity to cognitive decline/disruption over the lifespan (Levy et al., 2002; Guarino et al., 2019; Taconnat et al., 2022).

### 2.3 Sleep Assessments

Participants’ sleep health was assessed via the Pittsburgh Sleep Quality Index (PSQI) (Buysse et al., 1989), the Epworth Sleepiness Scale (Johns, 1991) and the Lausanne NoSAS (Marti-Soler et al., 2016). The latter two scales were not included in baseline assessments and therefore constitute less of the dataset than the PSQI (see **Table 1**). Higher scores on the latter scales indicate increased daytime sleepiness and an increased risk of sleep-disordered breathing. Due to the low predictive validity of a total PSQI score (Landry et al., 2015; Parsey et al., 2015), we chose to use several of its components: hours of sleep per night, waking in the night or early morning, and self-reported overall sleep quality. Note that higher ratings of the latter three PSQI components (scored 0 – 3) correspond to worse sleep.

**Table 1.** Participant characteristics, cognitive performance, sleep and affect scores

| Characteristics | N |  |  |
| --- | --- | --- | --- |
| Age (in years) | 700 | M(SD) | 51.17(5.47) |
| Sex | 700 | % Female | 61.86% |
| Education (in years) | 698 | M(SD) | 16.69(3.44) |
| Short Sleep Group | 217 | % Sleep Group | 31.04% |
| Medium Sleep Group | 361 | % Sleep Group | 51.65% |
| Long Sleep Group | 121 | % Sleep Group | 17.31% |
| **Executive Function** |  |  |  |
| Colour (RT) | 677 | M(SD) | 959.04(156.26) |
| Word (RT) | 677 | M(SD) | 1023.68(161.30) |
| Interference (RT) | 677 | M(SD) | 1458.81(321.80) |
| **Memory** |  |  |  |
| Cued (correct) | 681 | M(SD) | 7.01(1.43) |
| Free (correct) | 680 | M(SD) | 6.88(1.46) |
| Immediate (correct) | 680 | M(SD) | 6.59(1.32) |
| List (correct) | 678 | M(SD) | 17.06(1.27) |
| **Sleep** |  |  |  |
| Total Hours of Sleep | 699 | M(SD) | 6.76(0.98) |
| Sleep Quality | 698 | M(SD) | 1.07(0.76) |
| Waking in the Night/Morning | 700 | M(SD) | 2.13(1.00) |
| Epworth Scale | 257 | M(SD) | 5.81(4.12) |
| NoSAS Scale | 228 | M(SD) | 6.89(4.61) |
| **Affect** |  |  |  |
| Total CES-D Score | 700 | M(SD) | 16.12(5.50) |
| Total Apathy Score | 681 | M(SD) | 1.10(3.58) |

Note: Group means and standard deviations (M(SD)) for age (in years), years of education, Stroop colour, word and interference task reaction times (RT), number of correct cued, free, immediate, and list recall responses; total hours of sleep, self-reported sleep quality, prevalence of waking in the night or early morning, the Epworth Sleepiness Scale, the Lausanne NoSAS Scale, the Centre for Epidemiological Studies-Depression (CES-D) scale, and the PREVENT apathy scale. Note that we also present the % of females in our sample (N), and the percentage of participants belonging to the Short (i.e., < 6 hours), Medium (i.e., 6 – 7.99 hours) and Long (i.e., > 8 hours) sleep groups.

### 2.4 Depression Symptomology

The degree to which participants suffered from depressive symptoms was determined via the CES-D scale (Radloff, 1977). The CES-D is a 20-item depression symptom assessment with each item being scored on a scale 0-3 (i.e., total score from 0-60), and includes questions regarding participants’ feelings of loneliness and the degree to which they enjoy life. Higher scores indicate more symptoms.

### 2.5 Apathy Scores

Participants’ apathy was assessed via a 3-item apathy scale wherein participants reported whether they experienced “emotional blunting”, a “lack of initiative”, and/or a “lack of interest”. The frequency with which these symptoms occurred are summed to create a total score. Higher total scores are indicative of higher ratings of apathy. We chose to include a measure of apathy here because lack of interest/apathy has been defined as a core symptom of clinical depression (Blazer, 2003). Hence, apathy may indirectly mediate the association between depression symptoms and cognitive function (see Fishman et al., 2019).

### 2.6 Statistical Analyses

#### 2.6.1 Pre-processing

All data processing and subsequent analyses were performed in Stata SE 16.1. Prior to modelling, we assessed and processed responses to cued, free, immediate and list recall tasks; Stroop colour, word and interference RTs; sleep, and depression and apathy scores. Where appropriate, skewed (i.e., g_1_ > 1.0) data were log-transformed for normalisation. We note that apathy scores remained skewed following log-transformation and were subsequently z-transformed to minimise their lack of normality. All cognitive variables of interest and sleep scores were z-transformed to normalise scaling (see **Figure 1**).

**Figure 1.**
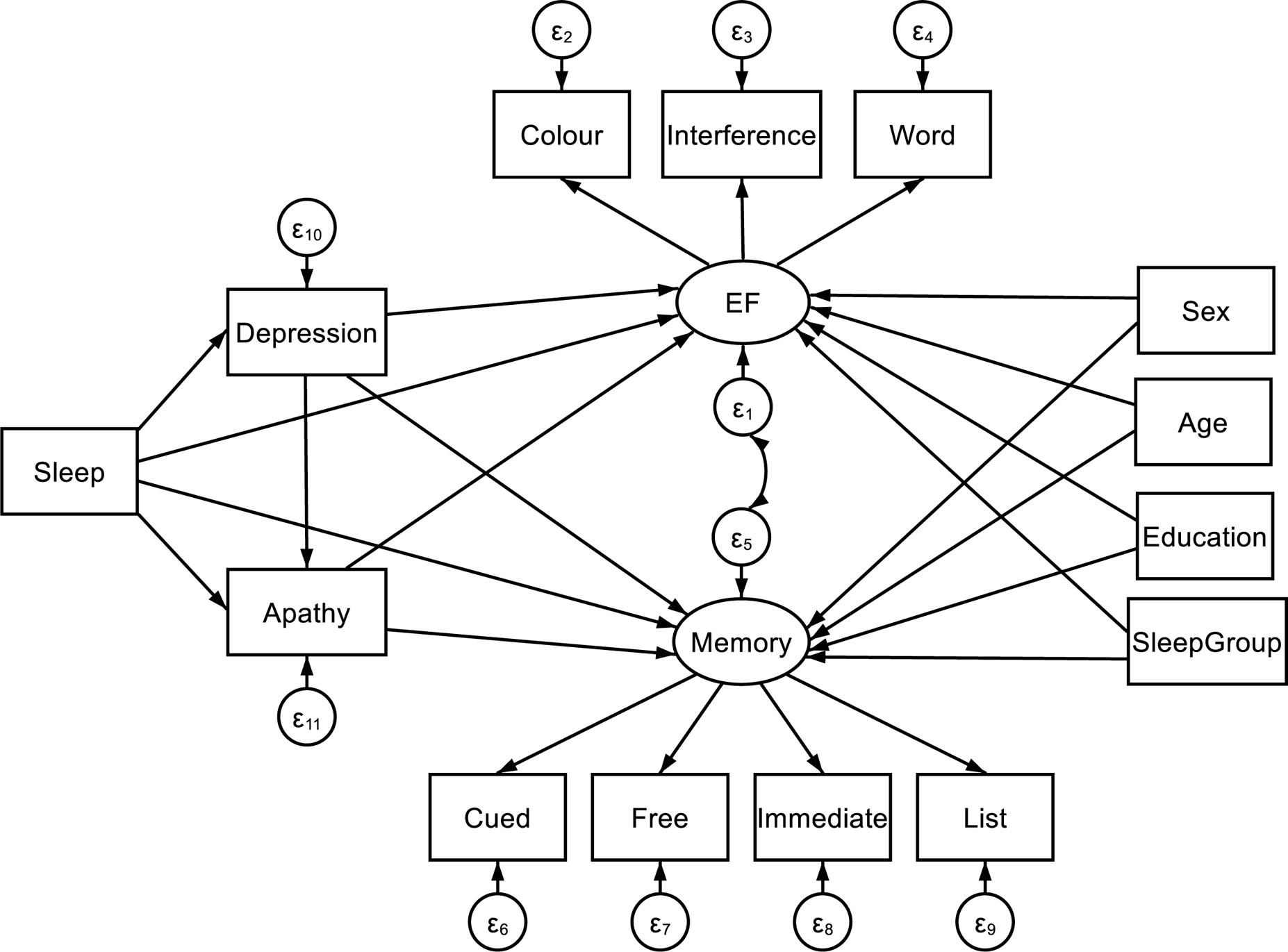
Structural equation model including predictor variables: Sleep, Depression (i.e., CES-D total score) and Apathy (i.e., apathy scale total score); mediator variables: Sex (i.e., male v. female), Age (in years), Education (i.e., total years of education), and SleepGroup (i.e., < 6 hours = Short; 6 – 7.99 hours = Medium; > 8 hours = Long). Note that for data visualisation purposes, Sleep has been inserted into the model as a single measurememnt; however, it is composed of separate measures of hours of sleep per night, waking in the night or early morning, self-reported overall sleep quality, the Epworth Sleepiness Scale, and the Lausanne NoSAS. Paths extend from these variables to two latent constructs: Executive Function (EF) (i.e., Stroop colour, word and interference task reaction times) and Memory (i.e., correct responses in cued, free, immediate and list recall tasks). The latent constructs are joined via a covariance link.

#### 2.6.2 Pairwise Correlations

Pairwise correlations were employed to explore any associations between participant age, sex, years of education, sleep (i.e., hours of sleep per night, waking in the night or early morning, self-reported overall sleep quality, Epworth Sleepiness Scale, Lausanne NoSAS), depression and anxiety scores, Stroop colour, word and interference task RTs, and correct responses to cued, free, immediate and list recall tasks. Correlations were Bonferroni corrected and associations were considered significant if p < 0.05.

#### 2.6.3 Structural Equation Model

We employed a structural equation model (SEM) to assess direct and indirect effects between sleep, depression, apathy and cognitive function. That is, we aimed to create a single model to assess a mechanistic pathway by which our predictor variables may influence cognition. Prior to creating our model, simple regressions of the variables of interest were performed to better inform direct and indirect model paths. The presented model was estimated using a maximum likelihood with missing values (MLMV) test. We report standardised coefficients and beta values. The MLMV method assumes joint normality and, if present, randomly occurring missing values. The resulting model contains the following variables.

Cognition was assessed via two latent constructs. First, Executive Function (EF) was composed of Stroop colour, word and interference task RTs (Periáñez et al., 2021). We then collated performance on the COGNITO tasks 8 and 17 into a Memory construct consisting of cued, free, immediate and list recall responses. A covariance link was applied between these constructs. Predictor variables included various indicators of sleep quality: TotalSleep (i.e., hours of sleep per night), WNEM (i.e., waking in the night or early morning), Quality (i.e., self-reported overall sleep quality), Epworth (i.e., the Epworth Sleepiness Scale), and NoSAS (i.e., Lausanne NoSAS). Depression and Apathy were included in our SEM as the total CES-D score, and total apathy score, respectively. Finally, Age and Education (i.e., years; continuous variables), SleepGroup (i.e., < 6 hours = 0, Short; 6 – 7.99 hours = 1, Medium; > 8 hours = 2, Long), as well as Sex (i.e., females = 1, males = 2) were entered into our SEM as covariates to control for any confounds. Effects were deemed significant when p < 0.05.

## 3.0 Results

On average, the included sample was 51.17 years old (SD = 5.47), comprised of mostly females (i.e., 62%), had completed 16.69 (SD = 3.44) years of education, and most slept between 6 and 8 hours a night (i.e., 52%) (see **Table 1**).

### 3.1 Pairwise Correlations

Initial pairwise correlations show no associations between predictor variables (i.e., indices of sleep, Apathy, Depression) and our chosen cognitive variables (rs < −0.002, ps > 0.99). We note, however, that Depression was related to TotalSleep, Quality, and WNEM (rs = −0.25, 0.36, 0.22, ps < 0.001), whereas Apathy was only related to Quality (r = 0.14, p = 0.047); Depression and Apathy were also related to each other (r = 0.23, p < 0.001). In addition, the PSQI measures used here were correlated (rs > −0.27, ps < 0.001) as well Stroop colour, word and interference task RTs (rs > 0.51, ps < 0.001), and the number of correct cued, free, immediate, and list recall responses (rs > 0.44, p < 0.001). Scores on the Epworth and NoSAS scales were not related to each other (r = 0.14, p > 0.99).

### 3.2 Structural Equation Model

#### 3.2.1 Regression Paths

As demonstrated in **Figure 1**, a direct path was extended from each predictor (i.e., Sleep including separate measures of TotalSleep, WNEM, Quality, Epworth, NoSAS, Depression, Apathy) and mediator (i.e., Sex, Age, Education, SleepGroup) variable to both cognitive latent constructs (i.e., EF and Memory). Links were also included between all sleep indices and Depression and Apathy, as well as between Depression and Apathy to assess the mediation of any relationship between our predictors and cognition.

#### 3.2.2 Estimation and Fit

Our model fit was deemed good according to accepted standards (e.g., Kline, 2016). Our model possesses a root mean squared error of approximation (RMSEA: differences between predicted and observed outcomes) = 0.025; the Tucker Lewis index (i.e., relative reduction in misfit per degree of freedom) = 0.982; the comparative fit index (CFI: metric of the model’s improvement from baseline to proposed iterations) = 0.988.

**Table 2** contains the SEM output for our model and demonstrates that worse sleep quality and fewer hours of sleep were associated with more depression symptoms (βs = 0.27, −0.10, ps < 0.01). Moreover, higher scores on the Epworth and NoSAS scales were associated with more apathy symptoms (βs = 0.16, 0.13, ps < 0.01) and depression and apathy symptoms were positively related (β = 0.20, p < 0.001). Neither Depression nor Apathy, nor any of the indices of sleep quality were related to memory performance (βs < −0.08, ps > 0.06); however, higher depression symptoms were found to be linked to worse executive function (i.e., longer RTs) (β = 0.12, p = 0.005) (see also **Figure 2**). Results demonstrated mediation of the effect of sleep on cognitive performance by depression symptoms. Indeed, β values regarding the relationship between sleep indices and executive function (β < −0.07) were attenuated by the effect of depression symptoms (β = 0.12). In terms of our covariates, we note that neither EF nor Memory were related to the sleep group to which participants belonged (i.e., Short: < 6 hours, Medium: 6 – 7.99 hours, Long: > 8 hours) (βs = −0.02, −0.11, ps > 0.21). Older age and less education were related to poorer executive function (βs = 0.27, −0.10, ps < 0.02), whereas being more highly educated and female were related to better memory performance (βs = 0.11, −0.27, ps < 0.01). The relationship between memory and participant age approached, but did not achieve conventional levels of statistical significance (β = −0.08, p = 0.06).

**Figure 2.**
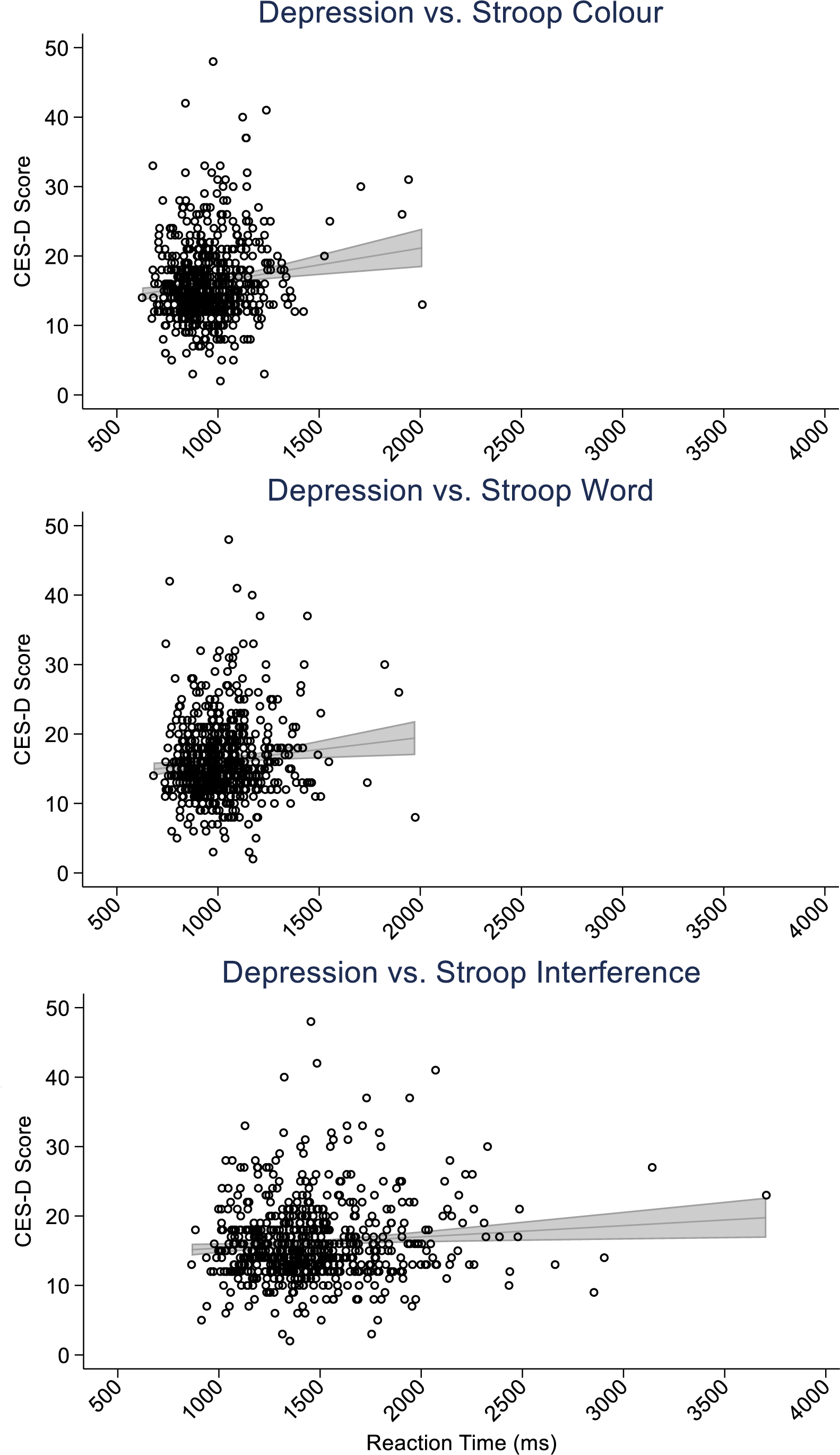
Scatter plots depicting the relationship between the Centre for Epidemiological Studies-Depression (CES-D) scale scores and Stroop colour (top), word (centre), and interference (bottom) task reaction times (ms). The panels include simple linear regressions accompanied by grey 95% confidence interval bands.

**Table 2.** Structural equation model output

| | Predictor | $\beta$ | SE | z | p | 95% CI | |
| --- | --- | --- | --- | --- | --- | --- | --- |
| Depression | WNEM | 0.06 | 0.04 | 1.46 | 0.14 | -0.02 | 0.14 |
|  | Quality | 0.27 | 0.04 | 6.31 | 0.000* | 0.19 | 0.36 |
|  | TotalSleep | -0.10 | 0.04 | -2.50 | 0.01* | -0.18 | -0.02 |
|  | Epworth | 0.05 | 0.06 | 0.76 | 0.45 | -0.08 | 0.18 |
|  | NoSAS | -0.04 | 0.05 | -0.94 | 0.35 | -0.14 | 0.05 |
| Apathy | Depression | 0.20 | 0.04 | 5.03 | 0.000* | 0.12 | 0.28 |
|  | WNEM | -0.01 | 0.04 | -0.16 | 0.87 | -0.09 | 0.08 |
|  | Quality | 0.05 | 0.05 | 0.93 | 0.35 | -0.05 | 0.14 |
|  | TotalSleep | -0.01 | 0.04 | -0.13 | 0.90 | -0.09 | 0.08 |
|  | Epworth | 0.16 | 0.06 | 2.58 | 0.01* | 0.04 | 0.29 |
|  | NoSAS | 0.13 | 0.05 | 2.64 | 0.01* | 0.03 | 0.22 |
| EF | Depression | 0.12 | 0.04 | 2.79 | 0.005* | 0.04 | 0.20 |
|  | Apathy | 0.07 | 0.04 | 1.73 | 0.08 | -0.01 | 0.16 |
|  | Sex | 0.03 | 0.07 | 0.47 | 0.64 | -0.10 | 0.16 |
|  | Age | 0.27 | 0.05 | 5.80 | 0.000* | 0.18 | 0.36 |
|  | Education | -0.10 | 0.04 | -2.37 | 0.02* | -0.18 | -0.02 |
|  | WNEM | 0.02 | 0.04 | 0.53 | 0.59 | -0.06 | 0.11 |
|  | Quality | -0.04 | 0.05 | -0.79 | 0.43 | -0.14 | 0.06 |
|  | TotalSleep | 0.04 | 0.09 | 0.42 | 0.67 | -0.14 | 0.22 |
|  | SleepGroup | -0.02 | 0.09 | -0.27 | 0.79 | -0.20 | 0.15 |
|  | Epworth | -0.07 | 0.07 | -1.00 | 0.32 | -0.21 | 0.07 |
|  | NoSAS | -0.03 | 0.10 | -0.33 | 0.74 | -0.23 | 0.16 |
| Memory | Depression | 0.01 | 0.04 | 0.28 | 0.78 | -0.07 | 0.09 |
|  | Apathy | -0.08 | 0.04 | -1.89 | 0.06 | -0.16 | 0.003 |
|  | Sex | -0.27 | 0.06 | -4.56 | 0.000* | -0.39 | -0.16 |
|  | Age | -0.08 | 0.04 | -1.88 | 0.06 | -0.17 | 0.004 |
|  | Education | 0.11 | 0.04 | 2.72 | 0.01* | 0.03 | 0.19 |
|  | WNEM | 0.04 | 0.04 | 0.94 | 0.35 | -0.04 | 0.13 |
|  | Quality | -0.08 | 0.05 | -1.68 | 0.09 | -0.18 | 0.01 |
|  | TotalSleep | 0.10 | 0.09 | 1.11 | 0.27 | -0.08 | 0.27 |
|  | SleepGroup | -0.11 | 0.09 | -1.26 | 0.21 | -0.28 | 0.06 |
|  | Epworth | 0.11 | 0.06 | 1.67 | 0.10 | -0.02 | 0.23 |
|  | NoSAS | -0.05 | 0.08 | -0.61 | 0.54 | -0.22 | 0.11 |

Note: The left most column indicates the variables of interest and includes: Depression (i.e., CES-D total scores), Apathy (i.e., apathy scale total scores), EF (i.e., Stroop colour, word and interference task reaction times in a latent construct) and Memory (i.e., a latent construct including correct responses to cued, free, immediate and list recall tasks). Predictor variables indicate those which have direct paths to the variable of interest. TotalSleep (i.e., hours of sleep per night), WNEM (i.e., waking in the night or early morning), Quality (i.e., self-reported overall sleep quality), Epworth (i.e., the Epworth Sleepiness Scale), NoSAS (i.e., Lausanne NoSAS), and SleepGroup (i.e., < 6 hours = Short; 6 – 7.99 hours = Medium; > 8 hours = Long). The output provides standardised beta ( $\beta$ ) values, standard error (SE), z scores, p values, and 95% confidence intervals (95% CI). For ease of interpretation, p values marked with \* indicate a statistically reliable association: $p < 0.05$ . N = 700.

## 4.0 Discussion

Our investigation sought to explore the association between sleep, depressive symptoms and cognition in healthy middle-aged adults. Below, we discuss the links between four confounding variables and the assessed cognitive constructs prior to explaining a model which demonstrates a direct link between depression symptoms and executive function.

### 4.1 Age, sex and years of education: mediators of memory and executive function

Our model showed no association between the sleep group to which individuals belonged and cognition. This is contrary to recent results presented by Ma and colleagues (2020) demonstrating an inverted-U relationship between sleep duration and the likelihood of global cognitive decline. We note that the authors had a considerably larger sample size (N = 20,065) and this is a likely explanation as to why this result is absent from our model. Moreover, we demonstrated that sex and education were related to memory and executive function. That is, being male (e.g., Voyer et al., 2021) and less educated (e.g., Murayama et al., 2013) was associated with fewer correct responses to memory tasks, whereas more education was related to improved executive function. Our memory composite comprised verbal memory subtasks tapping episodic memory, a domain typically demonstrating a performance advantage for women relative to men (Asperholm et al 2019), although inherent hormonal differences (i.e., estrogen concentrations) may modulate memory performance for females over time (Duarte-Guterman, 2015). Similarly, Staekenborg et al. (2020) and Han et al., (2023) indicate that increased cognitive reserve gained from more years of education may support memory and executive function in later life, while Joannette et al. (2020) show that educational attainment moderates the relationship between episodic memory and amyloid load. In contrast, being older was associated with worse performance on executive function. This is unsurprising as work has consistently demonstrated an association between increasing age and slowed RTs on executive functiontasks (Krivanek et al., 2021). What is more, detriments to global cognition related to age have been attributed to cortical thinning, demyelination, and brain volume loss (Blinkouskaya et al., 2021), as well as inefficient preparation of responses to stimuli (Williams et al., 2007; Hardwick et al., 2022). Accordingly, three of the confounding variables used here were related to cognition in a way that is aligned with the current corpus of literature.

### 4.2 Sleep may indirectly predict cognition via depression symptoms

Better sleep has been linked to improved mental health (Sadler et al., 2018), and has been shown to support cognition (Dzierzewski et al., 2018; Matricciani et al., 2019) and aide in the clearance of harmful Aβ protein (Xie et al., 2013; Tabuchi et al., 2015; Shokri-Kojori et al., 2018). In our model, however, none of the sleep measures investigated here were related to either cognitive construct. This may be an unexpected result given that cognitive dysfunction and insomnia share common neural mechanisms such as impaired functional connectivity and structural abnormalities within the amygdala, prefrontal cortex, anterior cingulate cortex and insula (e.g., Bagherzadheth-Azbari et al., 2019). Our null findings may be explained by the fact that the PREVENT cohort is composed of middle-, rather than old-aged, adults. This is notable because it is during this time in the lifespan where cognition is comparatively less vulnerable to insult (Diamond, 2013; Krivanek et al., 2021). However, even studies involving middle-aged individuals have demonstrated an association between sleep and impaired cognition (Ma et al., 2020), as well as Aβ concentrations (Sprecher et al., 2015). An alternative explanation may be that sleep is associated with cognition via some mediator(s).

Our model found that fewer hours of sleep and lower self-reported sleep quality were related to more symptoms of depression, and that higher depression symptoms and more daytime sleepiness and sleep apnoea likelihood (i.e., Epworth Sleepiness and NoSAS Scales) were related to participants’ total apathy score. We will address these results in turn. First, literature has previously demonstrated a link between sleep and depression (Scott et al., 2021; Joo et al., 2022). For example, a recent meta-analysis by Scott and colleagues (2021) provides evidence for a small-to-medium association between improved sleep quality and reduced symptoms of anxiety, stress, psychosis, and depression. Their analyses were conducted on studies from various countries and offers insight to the generalisability of this association across populations. In contrast, when Joo et al. (2022) assessed the relationship between each component of the PSQI and symptoms of depression, they found a dose-response relationship for each component of the index, except sleep duration. The link between sleep and depression has been explained biologically via increased activity in the amygdala (Yoo et al., 2007) and reduced functional connectivity between the amygdala and the prefrontal cortex (Motomura et al., 2013). What is more, one night of sleep deprivation has been linked with elevated sympathetic nervous system activity, increased heart rate variability and a subsequently diminished capacity to respond to emotional challenges (Sauvet et al., 2010; Zhong et al., 2005; Appelhans and Luecken, 2006; for review see Goldstein and Walker, 2014). Hence, that various measures of sleep were related to depression symptoms in our model was to be expected. Second, apathy affects various neurological outcomes and is common in individuals who present with symptoms of depression (Steffens et al., 2022); however, it is nosologically and neurobiologically distinct from depression (Tagariello et al., 2009). It is for this reason that we chose to include apathy in our model. We demonstrated that depression symptoms were unsurprisingly related to apathy scores and that apathy ratings were related to daytime sleepiness. Indeed, evidence has demonstrated that individuals with higher ratings of depression are less willing or likely to respond to rewards (Le Heron et al., 2018) and that resulting apathy symptoms are associated with alterations to frontoparietal executive networks (e.g., pre-frontal cortex and the anterior cingulate cortex; for review see Steffens et al., 2022). Previous work has shown that excessive sleepiness and sleep apnea disrupt normal activity within the pre-frontal cortex (e.g., Durning et al., 2014) and induce intermittent states of hypoxia (e.g., Bucks et al., 2013). These disruptions to neural activity and metabolism may be the mechanism(s) underlying how individuals develop/manifest feelings of apathy. When taken together, our results support literature demonstrating the links between less sleep, more depression symptoms and higher apathy present here.

Individuals with higher depressive and/or apathy symptoms often perform poorly on tests of executive function (e.g., Funes et al., 2018; McPherson et al., 2002). Here, our model demonstrated that depression, but not apathy, was associated with executive function. To understand these results, we considered the construction of our executive function latent construct. Rather than treating the Stroop colour, word, and interference tasks strictly as measures of stimulus processing and inhibition, respectively, we grouped them into one latent construct. This is because recent work from Periáñez et al. (2021) has demonstrated that performance on the Stroop colour, word, and interference tasks reflect visual search speed; Stroop colour and interference performance are indicative of working memory; and execution of the Stroop interference task is related to conflict monitoring. Accordingly, the ability to complete the various iterations of the Stroop task is dependent on a combination of several higher-order executive functions. Despite some evidence affirming the link between apathy and executive function, the literature is mixed. Tests of executive function and global cognition have yielded no reliable association with symptoms of apathy (Marin et al., 2003; Brodaty et al., 2010) and this can be explained by the different pathways by which apathy and executive function are mediated in the brain (see Gonsalves et al., 2020). On the other hand, depression has been shown to negatively alter performance on tasks which require top-down control. A recent investigation identified that symptoms of depression are related to higher cortical noise which negatively impacts executive performance (Yao et al., 2022). Although not directly assessed here, it is therefore possible that higher and increasingly inefficient frontoparietal brain activity associated with less sleep and higher depression symptoms (e.g., Steffens et al., 2022) underlies the detrimental relationship between depression and cognition.

Last, our model demonstrated neither depression symptoms nor apathy scores were related to Memory. The literature regarding these associations is mixed. For example, Fishman et al. (2019) demonstrated that stroke patients with elevated apathy ratings performed worse on free recall tasks, whereas depression symptoms did not elicit a similar result. Conversely, Szymkowicz et al. (2018) found that Parkinson’s disease patients with higher ratings of depression, but not apathy, performed worse on memory tasks. Our results do not seem to support these findings. However, it is worth noting that the relationships described above were found in individuals with psychiatric and/or physical co-morbidity, and in individuals who have been diagnosed with clinical depression and/or apathy disorders. Indeed, our results were obtained by modeling mostly cognitively and psychiatrically healthy individuals, and it may be the lack of relevant co-morbidity which spares any association between depression and/or apathy symptoms and memory performance.

### 4.3 Limitations, future directions and conclusions

We are aware that our study presents several limitations. First, our sample is middle-aged, cognitively healthy, well educated and ethnically homogenous. Therefore, it is unlikely that the present model can be generalised outside of this demographic. Future investigations should aim to explore a more diverse cohort in terms of age, socio-economic and cognitive status. Similarly, the presented model may serve as a launch point for investigations regarding the quantification of dementia likelihood via follow-up of this cohort, or further exploration of the biological mechanisms of the associations we present here; for example, via investigations of neurodegeneration and brain volume. It is possible our data show the effects of depression on both sleep and executive function which may be related to functional rather than organic mental disorder. Longitudinal data will be important to investigate this. Second, the CES-D and apathy scales used here are multi-component assessments of their respective constructs. That is, whether a specific sub-component of each scale drives the above interactions is not yet known. Moreover, as we incorporated self-report measures of sleep, future studies might benefit from incorporating objective measures to assess sleep duration/quality (e.g., actigraphy or polysomnography); this is especially relevant given data questioning the PSQI’s predictive validity for objective sleep duration and quality (Landry et al, 2015; Parsey et al, 2015). As well, Evangelista and colleagues (2021) have demonstrated the importance for objective measures of sleep. The authors found discrepancies related to objective (i.e., polysomnography, multiple sleep latency test) and subjective (i.e., Epworth Sleepiness Scale) reports of sleep. Third, we note that the amount of data available for the Epworth and NoSAS scales was much less than other sleep measures due to their late introduction into this protocol. It may be that the related results observed here were due to this discrepancy in Ns. Finally, although the observed link between executive function and depressive symptoms intimates a possible deleterious neurological/physiological cascade across frontoparietal regions, the data necessary to confirm or quantify this relationship are unavailable. Regardless of the aforementioned limitations, our study provides evidence for an association between sleep and cognitive function mediated by depression symptoms in a middle-aged population. These results are pertinent in so much as they will encourage further investigation of the indirect relationship between sleep and cognition and the relevance of this relationship for the development of dementia. Furthermore, this work may serve as a basis to further explore the potential nature and timing of any treatments to prevent or ameliorate the development of dementia in later life.

## Data Availability

The data and relevant codes will be made available upon reasonable request.

## 5.0 Acknowledgments

The authors would like to thank and acknowledge the PREVENT dementia programme study participants. The data collection was also supported by research facilities at West London NHS Trust, NHS Lothian, Oxford Health NHS Foundation trust and Cambridgeshire and Peterborough NHS Foundation trust and St James’s hospital, Dublin.

## 7.0 Ethics Statement

The data used here is secondary data where ethical approval has been approved by the source cohort. Multi-site ethical approval was granted for the PREVENT Dementia programme by the UK London-Camberwell St Giles National Health Service (NHS) Research Ethics Committee (REC reference: 12/LO/1023, IRAS project ID: 88938), which operates according to the Helsinki Declaration of 1975 (and as revised in 1983). Separate ethical applications for the Dublin site were submitted and given favourable opinions by the Trinity College School of Psychology Research Ethics Committee (SPREC022021-010), and the St James Hospital/Tallaght University Hospital Joint Research Ethics Committee.

## 9.0 Data Availability Statement

The data and relevant codes will be made available upon reasonable request.

## Footnotes

1 Results from this paper identified no relationship between amyloid burden and sleep quality in 143 elderly (i.e., 70-85 years of age) individuals.

